# PD-1/PD-L1 Blockade Combined with AbnobaViscum^®^ Therapy is Linked to Improved Survival in Advanced or Metastatic NSCLC Patients, an ESMO-GROW Related Real-World Data Registry Study

**DOI:** 10.1101/2024.10.24.24316043

**Authors:** Friedemann Schad, Anja Thronicke, Ralf-Dieter Hofheinz, Reinhild Klein, Patricia Grabowski, Shiao-Li Oei, Hannah Wüstefeld, Christian Grah

## Abstract

**Background:** Recent advancements in cancer treatment have shown the potential of PD-1/PD-L1 inhibitor (ICB) plus *Viscum album* L. (VA) therapy in improving survival rates for patients with advanced or metastasized non-small cell lung cancer (NSCLC). The objective of this study was to investigate factors associated with improved survival in NSCLC patients treated with a combination of ICB and VA.

**Methods:** Patients with advanced or metastasized NSCLC from the accredited national Network Oncology registry were included in the real-world data study adhering to ESMO-GROW criteria. The study was conducted with ethics approval. Survival and the impact on hazard were compared between patients receiving PD-1/PD-L1 inhibitor therapy alone versus combinational PD-1/PD-L1 inhibitors and abnobaViscum^®^ therapy. Adjusted multivariate Cox proportional hazard analysis was utilized to examine factors linked to survival.

**Results:** Enrolled patients (n = 300) had stage III or stage IV NSCLC, had a 1.19 male/female ratio and were 68 years old (median). Two hundred and twenty-two patients (74%) were in the control (CTRL, PD-1/PD-L1 inhibitor therapy) and seventy-eight patients (26%) in the combinational (COMB, PD-1/PD-L1 inhibitor plus abnobaViscum^®^ therapy) group. The three-year survival was significantly prolonged by 7 months when abnobaViscum^®^ therapy was added to the anti-PD-1/PD-L1 therapy (Comb: 13.8 months vs. Control: 6.8 months, p = 0.005). The three-year survival rate was 16.5% in the COMB group and two times higher than the three-year survival rate in the CTRL group (8.0%). Adjusted multivariable Cox regression analysis was performed for patients with PD-L1 positive (≥1%) NSCLC treated with a first-line PD-1 inhibitor and revealed that the addition of abnobaViscum^®^ therapy to anti-PD-1 significantly lowered the hazard of death by 75% in (aHR: 0.25; 95%CI: 0.11-0.60, p=0.002).

**Conclusions:** Our results indicate that addition of abnobaViscum^®^ therapy is significantly linked to enhanced survival in patients with advanced or metastasized NSCLC who are undergoing treatment with standard PD-1/PD-L1 inhibitor therapy irrespective of their age, tumor stage, ECOG status, surgery or radiation. The mechanisms could involve a synergistic modulation of the immune response, reduced primary PD-1/PD-L1 inhibitor resistance via immunogenic cell death and/or modification of the tumor microenvironment by combinational PD-1/PD-L1 inhibitor and abnobaViscum^®^ therapy. Our findings should be complemented with analyses of RCT or R-RCT.

**Trial registration:** The study was registered retrospectively (DRKS00013335).

## 1. Introduction

Lung cancer is the leading cause of cancer-related deaths and only one in ten patients with metastatic non-small cell lung cancer (NSCLC) survives 5 years or more. However, the advancements in immune checkpoint blockade (ICB) targeting PD-1 (programmed cell death protein 1) or PD-L1 (programmed death-ligand 1) receptors improved survival rates of patients with metastatic NSCLC (1). Combinations, timing and duration of currently approved PD-1 inhibitors (pembrolizumab, nivolumab) or PD-L1 inhibitors (durvualumab, atezolizumab) are constantly modified due to rapid advancements and developments in clinical trials (2-9). Current improvements involve the first-line application of anti-PD-1/PD-L1 therapy reflecting the race for a cure of such a destructive disease (4, 5, 9,).

*Viscum album* L. extracts (European white-berry mistletoe, VA) being applied in addition to PD-1/PD-L1 blockade in advanced or metastasized NSCLC patients were shown to be linked to improved overall survival (64). Clinical safety studies have found no safety issues with the useof VA in conjunction with anti-PD-1/PD-L1 therapy (13, 14, 40) and the national guideline for complementary therapies in oncological patients indicates there is no evidence of an increased rate of serious adverse events with the simultaneous use of immune checkpoint inhibitors and VA (46). The aim of this study was to assess the overall survival of patients with advanced or metastastatic NSCLC patients undergoing standard-oncological immunotherapy, both with and without abnobaViscum^®^ therapy. To achieve the study objectives, we chose RWD as our primary data source from registry-based data. This source was deemed particularly suitable due to its relevance and the ability to provide a comprehensive picture of real clinical practice. In recent years, the use of Real-World Data (RWD) has gained significance, particularly in evidence generation for clinical studies. This methodology facilitates the identification of relevant subgroups, such as elderly NSCLC patients with a performance status of ≥1 or those with multiple comorbidities, from real-world clinical settings. This approach generates targeted evidence essential for evaluating the effectiveness of ICB, both with and without abnobaViscum^®^ therapy.

## 2. Materials and Methods

### 2.1. Study Design

This study is a real-world data (RWD) analysis using information from a German Cancer Society accredited source, the oncological registry Network Oncology (NO) (31) in line with the ESMO – Guidance for Reporting Oncological real-Word Evidcence GROW criteria, see supplementary material. Enrolment of patients was from July 2015 to May 2023. Patients received abnobaViscum^®^ therapy in line with the summary of product characteristics (SmPC) (32) and at the discretion of the physician. PD-1/PD-L1 inhibitors were administered alone or in combination with VA according to standard clinical practice. The rationale for using VA therapy in patients in this study was to enhance survival, improve health-related quality of life, and alleviate cancer- and treatment-related symptoms. AbnobaViscum^®^ extracts were given to the patients at the physician’s discretion. The primary objective was to evaluate overall survival of patients with advanced or metastasized NSCLC receiving anti-PD-1/PD-L1 treatment with and without abnobaViscum^®^ therapy. The secondary outcome aimed to descriptively assess whether specific variables were linked to a reduced risk of death. The analysis included patients with advanced or metastatic NSCLC (UICC stage III-IV) who had received first-line immune checkpoint inhibitor therapy, with or without abnobaViscum^®^ therapy, and were registered in NO database. Additional inclusion criteria included patients aged 18 years and older, of any gender, who provided written consent. Demographic, diagnosis, tumor stage, treatment and survival as well as tumor board and last contact data were extracted from the NO registry. The use of VA extracts in an integrative oncological setting was documented, including start and end dates, dosages, data of host tree of the VA. Follow-up was routinely conducted six months after the initial diagnosis and annually in subsequent years. Loss to follow-up was defined as the absence of any follow-up visits.

### 2.2. Interdisciplinary Team

The multidisciplinary team of the presented study consisted of experts from various fields, including clinical practice, epidemiology, and biostatistics. This diversity of expertise was crucial in meeting the requirements of a successful RWD study according to the ESMO-GROW criteria. Through close collaboration, we ensured that all aspects of the study were comprehensively addressed.

### 2.3. Ethics issues

The study adheres to the principles outlined in the Declaration of Helsinki. Written informed consent was obtained from all patients prior to their enrollment. The study was approved by the ethics committee of the Medical Association Berlin (Eth-27/10).

### 2.4. Classification of Groups

Patients with NSCLC were categorized into histological subgroups non-squamous cell carcinoma, squamous cell carcinoma, or large cell carcinoma. Patients were then assigned to one of the two groups: 1) the CTRL group, which received only PD-1/PD-L1 inhibitors without VA therapy, or 2) the COMB group, which received PD-1/PD-L1 inhibitors along with additional abnobaViscum^®^ therapy. The allocation to treatment groups was non-randomized and determined by the physician, following detailed information and the patient’s decision regarding treatment options. The VA therapy included abnobaViscum^®^ (ABNOBA GmbH, Niefern-Öschelbronn) extracts and extracts from other producers (Helixor Heilmittel GmbH, Rosenfeld; Iscador AG, Arlesheim).

### 2.5. Determination of Sample Size

To determine the necessary sample size for a two-sided test with an80% power and a significance level of 5% using an allocation ratio of 0.2 (CTRL) to 0.8 (COMB) and an effect size of 0.6 (10, 33), a total of 219 patients would be required. This included 44 patients in the COMB and 175 patients in the CTRL group, in order to confirm a statistically significant treatment effect, as outlined by Schoenfeld et al. (34).

### 2.6. Statistical Methods

Continuous variables were summarized using the median and interquartile range (IQR), while categorical variables were reported as absolute and relative frequencies. Data distributions were visually inspected, and skewness was evaluated arithmetically. Patients with missing data were excluded from the analysis. Baseline characteristics and treatment regimens for both group were compared using the unpaired Student’s t-test for independent samples. Chi-square analysis was used for comparisons of categorial variables. All statistical tests were two-sided, and all analysis were exploratory in nature. Kaplan-Meier survival curves were generated for both groups CTRL and COMB group. Patient survival was calculated from the index date until the last recorded event including either the date of death, the last documented personal contact, the last interdisciplinary tumor board, or follow-up. For survival analyses, the index date was defined as the first date of start of anti-PD-1/PD-L1 therapy. Patients who had not died by the time of the analysis were censored. A year was defined as 365.25 days, and a month at 365.25/12 days.

To examine the influence of various factors on patient survival while minimizing potential confounding, we employed a multivariate stratified Cox proportional hazards model, adjusting for age, gender, tumor stage, ECOG performance status, PD-L1 status, and oncological treatment. Before conducting this analysis, we performed verification analyses to ensure that the proportional hazard assumptions were satisfied. All analyses were conducted using R-Studio version 2022.02.2 and R software version 4.1.2 (2021-11-01) “bird hippie”, which is a language and environment for statistical computing (35). For Kaplan-Meier survival analysis, and multivariate Cox proportional hazards analysis we utilized the R package ‘survival’ (version 3.5-5) (36). The ‘prodlim, package was used for implementing nonparametric estimators for censored event history (survival) analysis (version 2019.11.13) (37). To draw survival curves the package ‘survminer’ was used, version 0.4.9 (38). The statistical analyses in this study not only encompass the outcomes but also take into account the internal and external validity of the data. We conducted sensitivity analyses such as subgroup analyses to verify the robustness of our results, to reduce potential biases and to understand variations in the response.

## 3. Results

### 3.1. Baseline Characteristics

Three hundred (n = 300) patients with advanced or metastasized NSCLC who received PD-1/PD-L1 inhibitors as part of their standard of care being documented in the network oncology registry were included in the study. Out of the total, 222 patients (74%) were treated with immune checkpoint inhibitors alone without on the addition of abnobaViscum^®^ therapy (control group, CTRL) while 78 patients (26%) received PD-1/PD-L1 inhibition in combination with abnobaViscum^®^ therapy (combinational group, COMB) (see flowchart, Figure 1). In total, 300 patients were included for the Kaplan-Meier survival analysis and one hundred and twenty one patients were enrolled for the adjusted multivariate cox regression analysis. The latter group had a PD-L1 positive (≥1%) NSCLC and were treated with a first-line PD-1/PD-L1 inhibitor therapy only, see figure 1.

**Figure 1.**
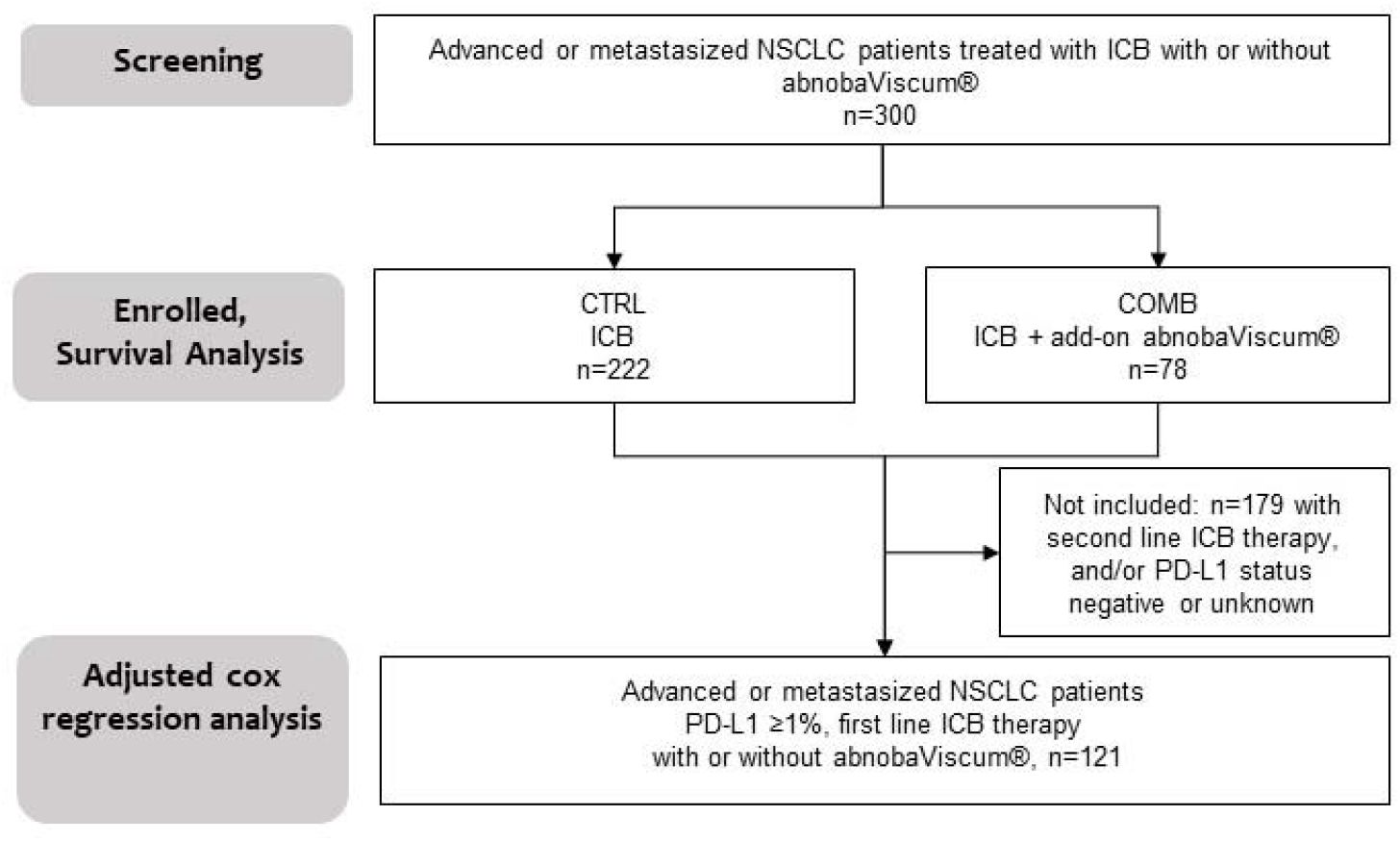
Study process flow. Patients with advanced or metastasized NSCLC who received PD-1/PD-L1 inhibitors, either with or without abnobaViscum^®^ therapy (n=300), CRTL, received PD-1/PD-L1 inhibitors and no abnobaViscum^®;^therapy; COMB, received PD-1/PD-L1 inhibitors in conjunction with abnobaViscum^®^ therapy; ICB, immune checkpoint blockade; n, number; abnobaViscum^®^, abnobaViscum^®^ therapy; PD-L1 ≥1%, ≥1% tumor proportion score of programmed death-ligand 1.

No significant differences were observed between the two groups regarding gender, histology, tumor stage, and surgery. The median age of the total cohort was 68 years (interquartile range 62-76). Participants from the COMB group were in median three years younger than participants from the CTRL group, the difference was not significant, see table 1. The sex ratio (male/female) was 1.19. The most common histological subtype of NSCLC was non-squamous cell carcinoma, accounting for 63% (n=189) of cases, followed by squamous cell carcinoma with 30% (n=90), as shown in table 1. In 7% of patients (n=21), the diagnosis of NSCLC was not further specified due to the nature of real-word data. In the COMB group, the percentage of patients with non-squamous cell carcinoma was slightly lower at 61.5% compared to 63.5% in the CRTL group, however, the differences between the groups were not statistically significant.

**Table 1.**
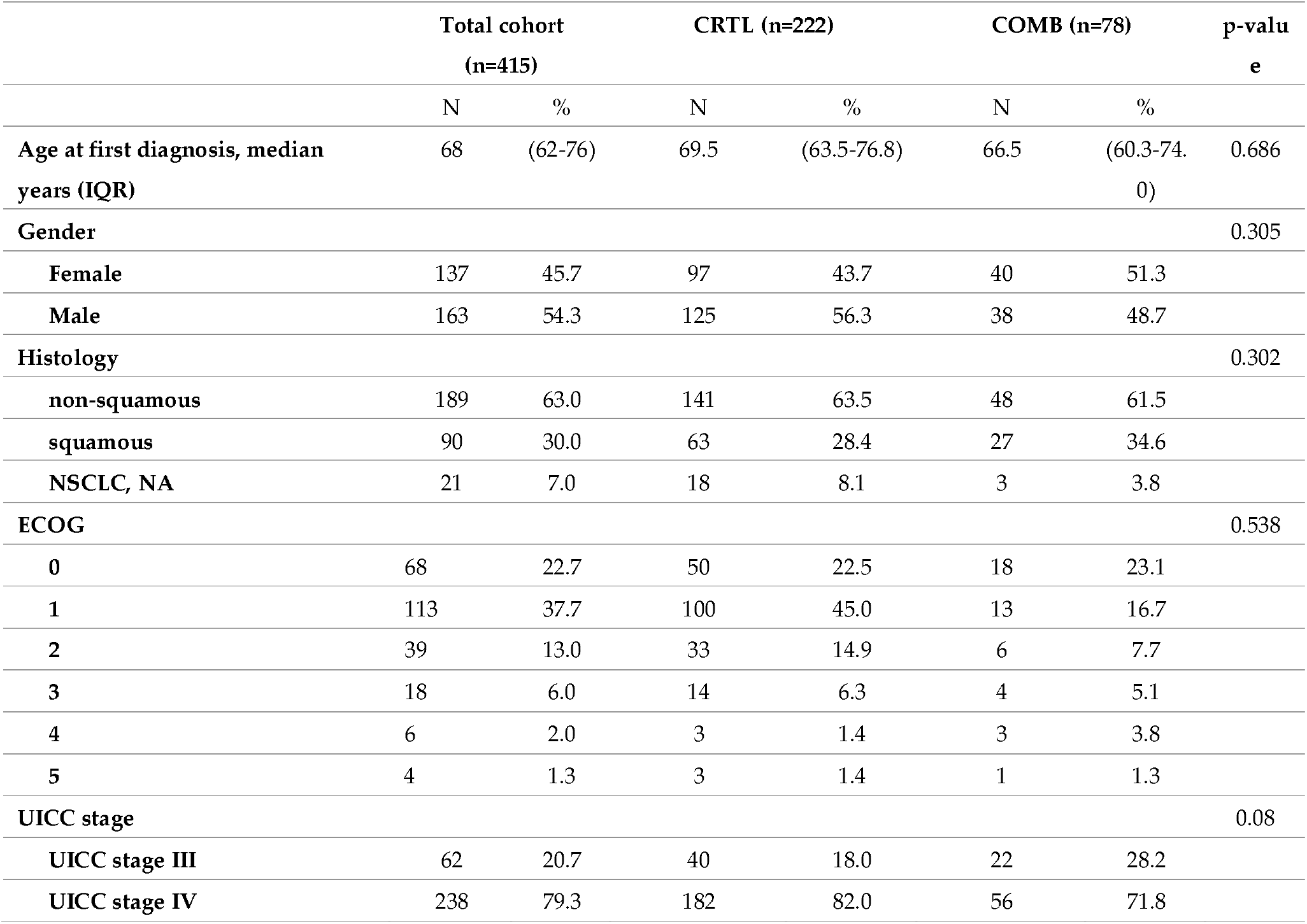
Characteristics of patients.

Patient characteristic, please note that the percentages of sub-variables may not sum to 100% due to rounding; IQR referes to the interquartile range; CRTL indicates patients treated with PD-1/PD-L1 inhibitors alone, while COMB referes to patients receiving PD-1/PD-L1 inhibitors in conjunction with VA therapy. UICC stands for Union International Contre le Cancer staging based on the UICC TNM classification system; ECOG represents the Eastern Cooperative Oncology Group.

### 3.2. Tumor Markers

No significant differences in molecular marker were noted between the two groups, except for PD-L1 status, as shown in table 2. In the CTRL-group, there was a 19.3% higher percentage of patients with a positive PD-L1 status and a 20.1% higher percentage with PD-L1 ≥50 TPS compared to the COMB group, and these differences were statistically significant. PD-L1 status was available for 82.3% (n=247) of the patients assessed, while the documentation of known BRAF mutations, EGFR (exon 18-21) mutations, ROS rearrangements, and ALK translocations varied from 52.3% to 100%, as detailed in table 2. Regarding stage IV NSCLC, the molecular status was recorded for 184 (61.3%) of the 238 patients.

**Table 2.** Molecular characteristics of patient’s non-small cell lung cancer.

|  | Total cohort (n=300) |  | CTRL (n=222) |  | COMB (n=78) |  | p-value |
| --- | --- | --- | --- | --- | --- | --- | --- |
|  | N | % | N | % | N | % |  |
| **PD-L1 status** |  |  |  |  |  |  | 0.007 |
| PD-L1 status, positive/known | 171/247 | 69.2 | 137/185 | 74.1 | 34/62 | 54.8 |  |
| PD-L1 status, positive TPS<br>≥50/known | 85/247 | 34.9 | 73 | 39.5 | 12 | 19.4 | 0.007 |
| **ALK translocation** |  |  |  |  |  |  | 1 |
| negative/known | 170/171 | 94.4 | 124/125 | 99.2 | 46/46 | 100 |  |
| **EGFR (Exon 18-21) mutation** |  |  |  |  |  |  | 0.070 |
| negative/known | 136/148 | 91.9 | 106/112 | 94.6 | 30/36 | 83.3 |  |
| **ROS1-rearrangement** |  |  |  |  |  |  | 0.596 |
| negative/known | 150/300 | 50.0 | 108/222 | 48.6 | 42/78 | 53.8 |  |
| **BRAF V600E-mutation** |  |  |  |  |  |  | 0.724 |
| negative/known | 154/157 | 98.1 | 114/117 | 97.4 | 40/40 | 100 |  |

Molecular characteristics of patients with NSCLC, CRTL refers to patients receiving PD-1/PD-L1 inhibitors alone, while COMB denotes patients treated with PD-1/PD-L1 inhibitors plus VA. ALK, anaplastic lymphoma kinase; BRAF, B-rapidly accelerated fibrosarcoma; EGFR, epidermal growth factor receptor; PD-L1 programmed death ligand 1; ROS1, receptor tyrosine kinase encoded by the ROS1 gene; TPS, tumor proportion score.

### 3.3. Oncological Treatment

Almost 60 percent of all enrolled patients received a first-line PD-1/PD-L1 therapy. PD-1 inhibitors were the most group (92%) compared to PD-L1 inhibitors (7%), see table 3. There was no significant difference in PD-1 or PD-L1 inhibitor treatment between the two groups. 13.6% more patients from the CTRL group received first-line treatment, a difference which was close to significance. Eleven percent of enrolled patients received a radiation of the lung and nine percent a radiation of the brain. Eleven percent of the patients also received a surgery and four percent a chemotherapy. While 9.3% more patients of the COMB group received surgery compared to CTRL (p=003) no significant differences as to radiation or chemotherapy between the two groups were observed.

**Table 3.** Characterization of antineoplastic therapy.

|  | Total cohort (n=300) |  | CTRL (n=222) |  | COMB (n=78) |  | p-value |
| --- | --- | --- | --- | --- | --- | --- | --- |
|  | N | % | N | % | N | % |  |
| **Radiation, bone** | 25 | 8.3 | 17 | 7.7 | 8 | 10.3 | 0.64 |
| **Radiation, brain** | 26 | 8.7 | 17 | 7.7 | 9 | 11.5 | 0.42 |
| **Radiation, primary tumour** | 33 | 11.0 | 21 | 9.5 | 12 | 15.4 | 0.22 |
| **Radiation, abdomen** | 1 | 0.3 | 1 | 0.5 | 0 | 0 | 1 |
| **Surgery** | 33 | 11.0 | 19 | 8.6 | 14 | 17.9 | 0.03 |
| **Chemotherapy** | 12 | 4.0 | 12 | 5.4 | 0 | 0 | 0.08 |
| **First-line immunotherapy** | 173 | 57.7 | 136 | 61.3 | 37 | 47.7 | 0.05 |
| **PD-L1/PD-1/CTL-A4 inhibitors** |  |  |  |  |  |  | 0.48 |
| PD-L1 inhibitors | 22 | 7.3 | 15 | 6.8 | 7 | 9.0 |  |
| PD-1 inhibitors | 275 | 91.7 | 204 | 91.9 | 71 | 91.0 |  |
| CTL-A4 inhibitor | 3 | 1.0 | 3 | 1.4 | 0 | 0 |  |

Oncological therapy; n, number of patients; %, percent. CTL-A4, cytotoxic T-lymphocyte antigen 4; PD-L1, programmed death ligand; PD-1, programmed cell death protein 1; CRTL, patients receiving PD-1/PD-L1 inhibitors without VA therapy; COMB, patients receiving PD-1/PD-L1 inhibitors plus VA therapy

### 3.4. Characterization of Combinational PD-1/PD-L1 Inhibitor and VA Therapy

The median duration of PD-1/PD-L1 therapy was 135 days (IQR 48-242 days) or 4.42 months (IQR 1.6-7.9 months) while VA therapy lasted in median 242 days (IQR 48-464 days) or 7,9 months (IQR 1,6-15,2 months).

Among the patients who received PD-1 inhibitors (either pembrolizuamb or nivolumab) 50.7% received concomitant intravenous, 36% concomitant subcutaneous, and 13.3% concomitant intratumoral abnobaViscum^®^ therapy, see figure 2.

**Figure 2.**
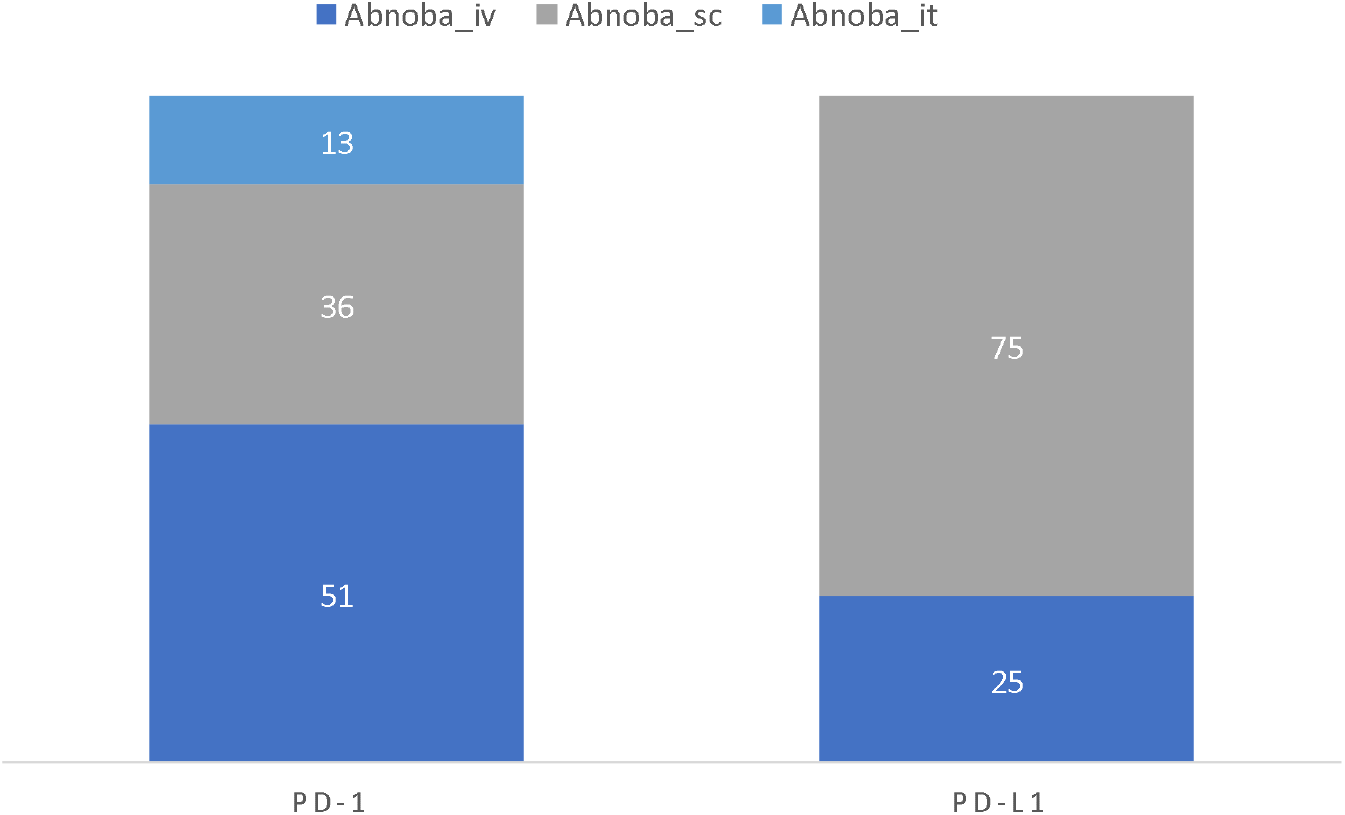
Characterization of combinational PD-1/PD-L1 inhibitor and abnobaViscum® therapy; Abnoba, abnobaViscum^®^ therapy; iv, intravenous; sc, subcutaneous; it, intratumoral.

Among the patients who received PD-L1 inhibitors (either atezolizumab or durvalumab) 25% received concomitant intravenous and 75% concomitant subcutaneous abnobaViscum^®^ therapy, see figure 2. In most cases the intravenous, subcutaneous or intratumoral abnobaViscum^®^ therapy was abnobaViscum^®^ fraxini (fraxini = ash tree) VA extract being applied to the patients at different doses and different application forms, see table 4. From all patients 37.2% patients received intravenous abnobaViscum^®^ fraxini with VA from other producers, see table 4. Combinations with VA from other producers were in cases of subcutaneous abnobaViscum^®^ fraxini application 19.2% and in intratumoral application 7.7% of all cases, see table 4. Other abnobaViscum^®^ applications included subcutaneous abnobaViscum^®^ abietis (fir tree), amygdali (almond tree) or quercus (oak tree) extracts that were combined VA from other producers (4.2%).

**Table 4.**
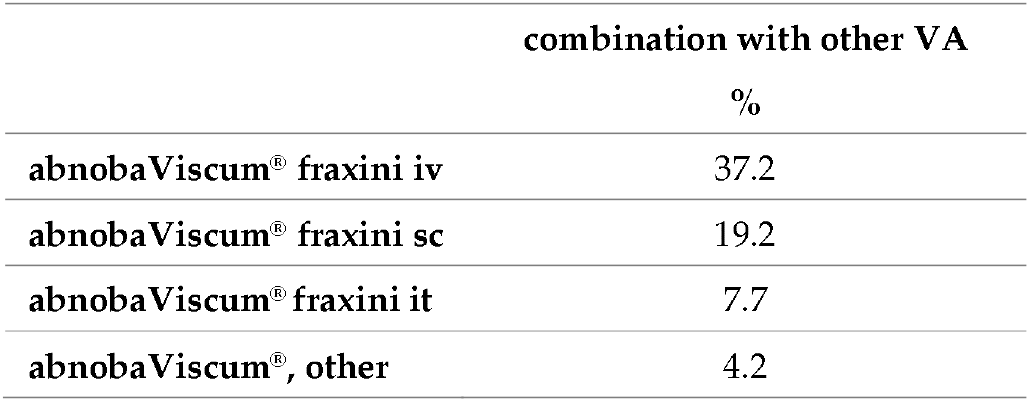
Application and combination forms of add-on abnobaViscum^®^ therapy . Characterization of VA therapy; VA, Viscum album L.; NA, not applicable; n, number; %, percent; iv, intravenous; sc, subcutaneous; it, intratumoral; other, other host tree.

### 3.5. Overall Survival of Advanced or Metastasized NSCLC Patients treated with PD-1/PD-L1 Inhibitors Plus Add-on AbnobaViscum^®^

A Kaplan-Meier survival analysis was performed for three hundred patients. Regarding the three-year survival, the COMB treatment (PD-1/PD-L1 inhibitors + abnobaViscum^®^ therapy) demonstrated a survival advantage compared to the CTRL group (PD-1/PD-L1 inhibitors without VA), as illustrated in figure 3. The median survival in the COMB group was 13.8 months (95%CI: 9.2 – 22 months), which is seven months longer than the median survival in the CTRL group which was 6.8 months (95%CI: 4.9 – 10.4 months). The log-rank test showed a significant difference (X^2^ = 7.9, p=0.005), as detailed in table 5. Out of the 300 patients, 187 patients (62.3%) died during the total observational period, with 62.8% (n = 49) in the COMB group and 62.2% (n = 138) in the CTRL group. The three-year survival rate was 16.5% in the COMB group, which is twice as high as the 8% three-year survival rate in the CTRL group.

**Table 5.** Median overall survival in patients with advanced or metastasized NSCLC in relation to treatment.

|  | N | Events | Median [months] | 95% CI [months] |
| --- | --- | --- | --- | --- |
| NSCLC, CTRL | 222 | 138 | 6.8 | 4.9 – 10.4 |
| NSCLC, COMB | 78 | 49 | 13.8 | 9.2 – 22.0 |
| Log rank test $X^2=7.9$, $p=0.005$ | | | | |

**Figure 3.**
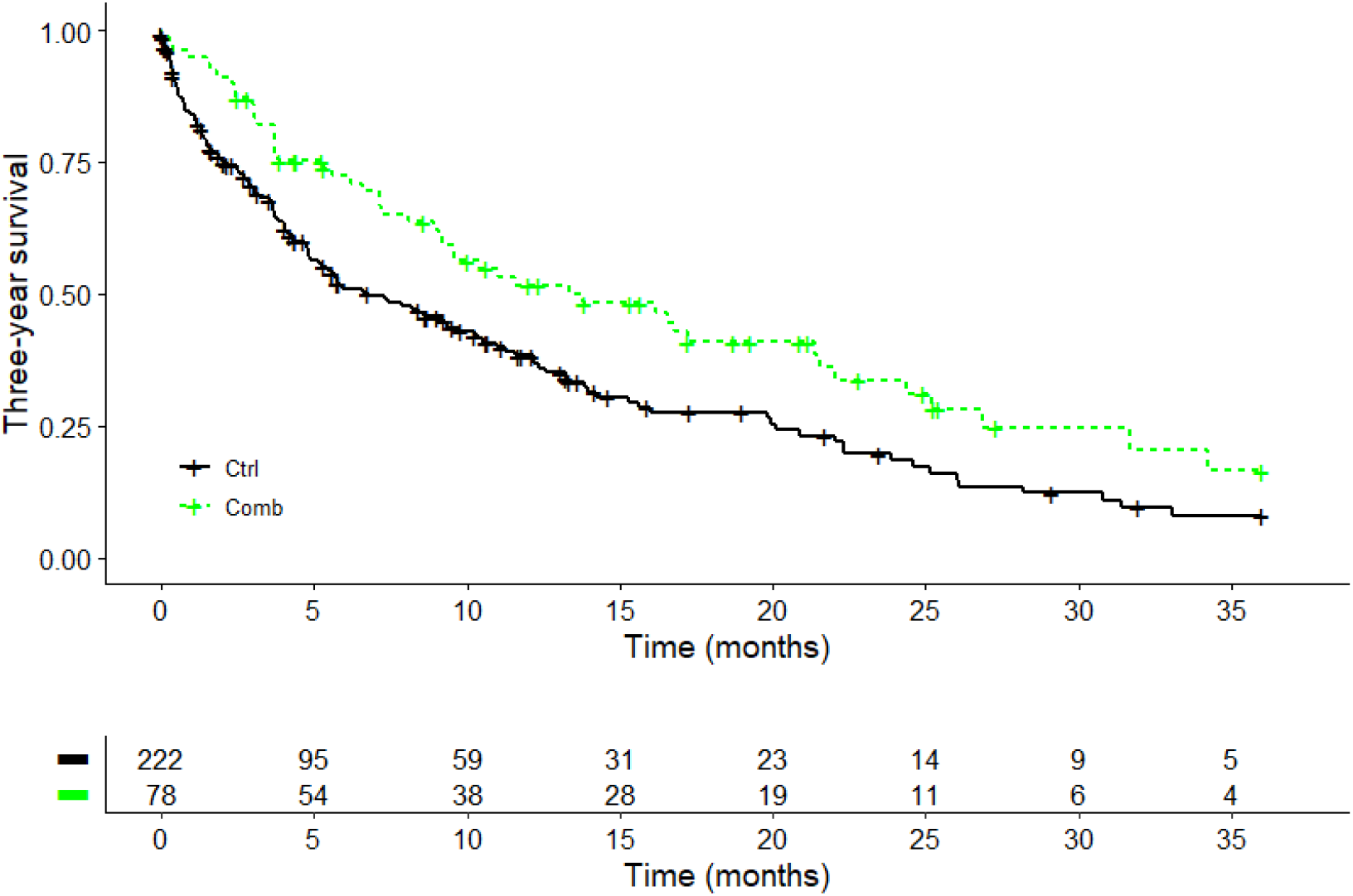
Kaplan–Meier survival curves displaying three-year survival. according to treatment in advanced or metastasized NSCLC (n=300); Log-rank test: X^2^ = 7.9, p=0.005; Ctrl, PD-1/PD-L1 inhibitors; Comb, PD-1/PD-L1 inhibitors + abnobaViscum^®^ therapy.

### 3.6. Add-on AbnobaViscum^®^ Therapy is Associated with Reduced Hazard of Death in PD-L1 Positive (≥1%) NSCLC Patients Treated with First-Line PD-1 Inhibitors

The adjusted multivariate Cox proportional hazard analysis on PD-L1 positive (≥1%) patients with advanced or metastasized NSCLC receiving first-line anti-PD-1 treatment indicated a statistically significant 75% reduction in the hazard of death (adjusted hazard ratio – aHR: 0.25, 95%CI: 0.11– 0.60, p = 0.002) when abnobaViscum^®^ therapy was added, as outlined in table 6. This effect was independent of gender, age, ECOG performance status, tumor stage, surgery, radiation, or PD-L1 TPS.

**Table 6.** Factors associated with hazard of death. Multivariate cox proportional analysis of factors linked to hazard of death in patients with advanced or metastasized PD-L1 positive NSCLC receving first-line PD-1/PD-L1 inhibitors, n=110, number of events 59, 11 observations deleted due to missing data; aHR, adjusted hazard ratio of death; VA, abnobaViscum^®^.; UICC, UICC, union international contre le cancer; PD-L1, programmed death-ligand 1; TPS, tumor proportion score, Score (logrank) test = 21.45 on 8 df, p=0.006^®.^

|  | aHR | (95% CI) | p-Value |
| --- | --- | --- | --- |
| abnobaViscum® therapy vs. non-VA | 0.25 | 0.11 - 0.60 | 0.002** |
| Age | 1.00 | 0.98 - 1.04 | 0.551 |
| Female gender vs. male gender | 0.78 | 0.42 - 1.45 | 0.430 |
| ECOG | 1.31 | 1.05 - 1.64 | 0.017* |
| Surgery vs. no surgery | 0.84 | 1.19 - 0.33 | 0.726 |
| UICC stage IV vs. stage III | 1.84 | 0.65 - 5.23 | 0.250 |
| Brain radiation vs. no radiation | 0.99 | 0.39 - 2.47 | 0.975 |
| PD-L1 TPS ≥50 | 0.60 | 1.66 - 0.34 | 0.090 |

## 4. Discussion

In this RWD study we assessed the effectiveness of PD-1/PD-L1 inhibitor therapy when combined with abnobaViscum^®^ therapy in cancer patients. Our results indicate that patients with advanced or metastasized NSCLC who received PD-1/PD-L1 inhibitors in combination with abnobaViscum^®^ therapy experienced improved survival compared to those patients receiving PD-1/PD-L1 inhibitors alone. Furthermore, patients with a PD-L1 positive (≥1%) NSCLC tumor treated with first-line PD-1 inhibitors in combination with abnobaViscum^®^ therapy had a better survival compared to patients treated with PD-1 inhibitors without add-on abnobaViscum^®^ therapy regardless of gender, age, tumor stage, ECOG performance score, oncological treatment, or PD-L1 TPS.

Findings from recent real-world data studies revealed that the combination of immune checkpoint blockade and VA therapy in advanced or metastasized NSCLC patients was associated with improved overall survival (64) and showed no safety concerns for VA (13, 14, 40,47). First prospective data results of PD-1/PD-L1 inhibitor in combination with VA therapy (Phoenix-3 study) in advanced or metastatic NSCLC patients confirm that the adverse event rates in patients receiving either PD-1/PD-L1 inhibitors alone or PD-1/PD-L1 inhibitors with mistletoe did not significantly differ (47). Recently, a review was published by the British Society for Integrative Oncology and Imperial College London on integrative oncology therapies that support immune checkpoint inhibitor therapy in solid tumours. The review concludes that the current data on immunotherapy - mistletoe combinations are still limited, however they consistently show no safety signal and are in accordance with clinical experiences of the authors (48). In accordance, the national guideline for complementary therapies in oncological patients indicated that there is no evidence of an increased rate of serious adverse events with the simultaneous use of immune checkpoint inhibitors and VA (46).

Clinical evidence is increasingly demonstrating the survival benefits of VA in cancer patients (35). Multiple systematic reviews, meta-analyses, and both clinical and real-word data studies suggest a positive impact of VA extracts on survival outcomes (10-12, 15-19, 65). The cause for the additive effect of VA on survival in NSCLC patients treated with PD-1/PD-L1 inhibitors in this study remains open. At least, VA seem not to interfere with the expression of PD-ligands on cancer cells in vitro (49). If they may interact with the

PD-1 receptor on immunocompetent cells is not yet known. However, VA have been shown to stimulate γβ T cells (50), which have strong antitumor effects in several tumors including NSCLC; these cells are also targeted by PD-1/PD-L1 inhibitors (51-53). This could explain why VA enhance the effect of PD-1/PD-L1 inhibitor therapy. On the other hand, blocking PD-L1 on cancer cells or PD-1 on T cells by specific antibodies may create a microenvironment, which allows VA to exert their anticancer effect by other immunological mechanisms. Thus, it has been indicated that various antigens found in VA extracts, such as mistletoe lectin and viscotoxins, are potent modulators of several cell types within both the innate and adaptive immune systems. These antigens interact with toll-like receptors on antigen-presenting cells, as well as with macrophages, natural killer cells, neutrophils, eosinophils, and T and B cells (54, 58). It is important to outline that VA rather modulate and do not activate or inhibit immunocompetent cells, which explains their good tolerability and the low incidence of severe side effects.

It has been shown that incidence of primary resistance to first- or second line PD-1/PD-L1 inhibitors ranging from 21% to 44% in NSCLC patients could be lowered to 7% to 11% when this therapy was combined with chemotherapy (57). This effect may be explained by the immunogenic cell death (ICD) induced by several chemotherapeutics such as cyclophosphamide and oxaliplatin (58) making the tumor cells more visible to the immune system so that primary resistance to PD-1/PD-L1 inhibitors can be overcome (63). In relation to that, VA extracts enhance the maturation of dendritic cells (54), are involved in the activation of immune cells (NK-cells, macrophages, dendritic cells (54, 58, 56), in cytokine (IL-1ß, IL-6, TNF-alpha) production (59, 60) as well as in antiangiogenic (61) and pro-apoptotic processes (62) – altogether mechanisms that qualify VA for a role in inducing ICD. During ICD the dying tumor cells release damage-associated molecular patterns (DAMPs) activating dendritic cells which initiate and manage an effective immune response against the tumor cells (63). Thus, add-on VA in the present study may be involved in ICD and the overcoming of primary resistance to applied immune checkpoint inhibitors in the present study resulting in longer overall survival in the NSCLC group with the combined treatment. Last, but not least, improved survival in the COMB group could be explained by a modified tumor microenvironment (TME). Immune checkpoint inhibitors can reduce the immunosuppressive signals in the tumor microenvironment, allowing immune cells to infiltrate and attack the tumor more effectively. VA extracts have been shown to inhibit angiogenesis and thus change the TME for a reduced growth of tumor cells and higher accessibility to immune cells.

### Limitations and Strength

The non-randomized character of the present real-world data study limits our results. However, the groups compared were well-balanced, minimizing the risk of comparing heterogeneous patient populations in terms of tumor type, stage of the disease,or oncological therapy. Additionally, potential biases were addressed using multivariable logistic regression methods in the survival analyses to account for confounding factors. A key strength of this real-world data study is that it reflects the acutal use of PD-1/PD-L1 inhibitor therapy in NSCLC patients and is the first to demonstrate a positive association between combined PD-1/PD-L1 inhibitor/abnobaViscum^®^ therapy and improved survival. Further research with larger and more diverse patient populations is needed to validate these results and gain a deeper understanding of the effects of combined PD-1/PD-L1 inhibitor and VA therapy in lung cancer treatment.

## 5. Conclusions

Our findings indicate that the addition of abnobaViscum^®^ therapy is significantly associated with enhanced survival in patients with advanced or metastatatic NSCLC receiving standard PD-1/PD-L1 inhibitor therapy, regardless of age, gender, metastatic status, or oncological treatment regimen. While these results highlight the clinical impact of add-on VA therapy, they should be further complemented with analyses of RCT or R-RCT.

Author contributions: FS and AT made significant contributions to the study design and planning, collected, interpreted and analysed the data, drafted the manuscript, critically revised it, and gave final approval for the version to be published. CG, RDH, RK, SLO, HW and PG contributed substantially to data interpretation, critically revised the manuscript, and gave final approval for the version to be published.

## Data Availability

All data produced in the present work are contained in the manuscript

## Institutional Review Board Statement

The study was carried out in compliance with the Declaration of Helsinki and received approval from the Ethics Committee of the Medical Association Berlin (Ethik-Kommission der Ärztekammer Berlin, protocol code Eth-27/10.

## Informed Consent Statement

Informed consent was obtained from all subjects in the study.

## Consent for publication

All authors consented to this manuscript’s publication.

## Data availabilty statement

All relevant data are included in this manuscript.

## Acknowledgments

We would like to express our gratidude to the collegues from the hospital Havelhöhe and the Research Institute Havelhöhe for their contributions to this work.

## References

1. Cancer.net, statistics, non-small cell lung cancer. Accessed 20.07.2023. https://www.cancer.net/cancer-types/lung-cancer-non-small-cell/statistics

2. Langer CJ. Emerging immunotherapies in the treatment of non-small cell lung cancer (NSCLC): the role of immune checkpoint inhibitors. Am J Clin Oncol. 2015 Aug;38(4):422–30. doi: 10.1097/COC.0000000000000059. PMID: 24685885.

3. Waldman, A.D., Fritz, J.M. & Lenardo, M.J. A guide to cancer immunotherapy: from T cell basic science to clinical practice. Nat Rev Immunol 20, 651–668 (2020). 10.1038/s41577-020-0306-5

4. EMA. EPAR Keytruda. https://www.ema.europa.eu/en/medicines/human/EPAR/keytruda

5. Bristol-Myers_Squibb. Positive Opinion Recommendation EMA. Accessed 21.07.2023 https://news.bms.com/news/details/2020/Bristol-Myers-Squibb-Receives-Positive-CHMP-Opinion-Recommending-Approval-of-Opdivo-nivolumab-Plus-Yervoy-ipilimumab-Combined-with-Two-Cycles-of-Chemotherapy-as-First-Line-Treatment-of-Metastatic-Non-Small-Cell-Lung-Cancer/default.aspx

6. EMA. EPAR Opdivo. Accessed 20.07.2023. https://www.ema.europa.eu/en/medicines/human/EPAR/opdivo

7. EMA. EPAR Imfinzi. Accessed 21.07.2023. https://www.ema.europa.eu/en/medicines/human/EPAR/imfinzi

8. EMA. EPAR Libtay. Accessed 21.07.2023. https://www.ema.europa.eu/en/medicines/human/EPAR/libtayo

9. EMA. EPAR Tecentriq. Accessed 21.07.2023. https://www.ema.europa.eu/en/medicines/human/EPAR/tecentriq

10. Schad F, Thronicke A, Steele ML, Merkle A, Matthes B, Grah C, Matthes H. Overall survival of stage IV non-small cell lung cancer patients treated with Viscum album L. in addition to chemotherapy, a real-world observational multicenter analysis. PLoS One. 2018 Aug 27;13(8):e0203058. doi: 10.1371/journal.pone.0203058. Erratum in: PLoS One. 2022 Aug 16;17(8):e0273387. PMID: 30148853; PMCID: PMC6110500.

11. Bar-Sela G, Wollner M, Hammer L, Agbarya A, Dudnik E, Haim N. Mistletoe as complementary treatment in patients with advanced non-small-cell lung cancer treated with carboplatin-based combinations: a randomised phase II study. Eur J Cancer. 2013;49:1058–1064

12. Salzer G, Danmayr E, Wutzholfer F, et al.: [Adjuvant Iscador® treatment of non-small cell bronchial carcinoma. Results of a randomized study]. Dtsch Z Onkol 23 (4): 93–8, 1991.

13. Thronicke A, Steele ML, Grah C, Matthes B, Schad F. Clinical safety of combined therapy of immune checkpoint inhibitors and Viscum album L. therapy in patients with advanced or metastatic cancer. BMC Complement Altern Med. 2017 Dec 13;17(1):534

14. Thronicke A, Oei SL, Grah C, Matthes B, Schad F. Nivolumab-induced toxicity profile in patients with advanced or metastasized lung cancer treated with Viscum album L. extracts. Abstract. German Cancer Congress, 2018

15. Loef M, Walach H. Survival of Cancer Patients Treated with Non-Fermented Mistletoe Extract: A Systematic Review and Meta-Analysis. Integr Cancer Ther. 2022 Jan-Dec;21:15347354221133561. doi: 10.1177/15347354221133561. PMID: 36324298; PMCID: PMC9634211.

16. Loef M, Walach H. Survival of Cancer Patients Treated with Non-Fermented Mistletoe Extract: A Systematic Review and Meta-Analysis. Integr Cancer Ther. 2022 Jan-Dec;21:15347354221133561. doi: 10.1177/15347354221133561. PMID: 36324298; PMCID: PMC9634211.

17. Tröger W, Galun D, Reif M, Schumann A, Stanković N, Milićević M. Viscum album [L.] extract therapy in patients with locally advanced or metastatic pancreatic cancer: a randomised clinical trial on overall survival. Eur J Cancer. 2013 Dec;49(18):3788–97. doi: 10.1016/j.ejca.2013.06.043. Epub 2013 Jul 24. PMID: 23890767.

18. Axtner J, Steele M, Kröz M, Spahn G, Matthes H, Schad F. Health services research of integrative oncology in palliative care of patients with advanced pancreatic cancer. BMC Cancer. 2016;16:579. Published 2016 Aug 2. doi:10.1186/s12885-016-2594-5

19. Loef M, Walach H. Quality of life in cancer patients treated with mistletoe: a systematic review and meta-analysis. BMC Complement Med Ther. 2020 Jul 20;20(1):227. doi: 10.1186/s12906-020-03013-3. PMID: 32690087; PMCID: PMC7370416.

20. Pelzer F, Loef M, Martin DD, Baumgartner S. Cancer-related fatigue in patients treated with mistletoe extracts: a systematic review and meta-analysis. Support Care Cancer. 2022 Aug;30(8):6405–6418. doi: 10.1007/s00520-022-06921-x. Epub 2022 Mar 3. PMID: 35239008; PMCID: PMC9213316.

21. Kienle GS, Kiene H. Review article: Influence of Viscum album L (European mistletoe) extracts on quality of life in cancer patients: a systematic review of controlled clinical studies. Integr Cancer Ther. 2010 Jun;9(2):142–57. doi: 10.1177/1534735410369673. Epub 2010 May 18. PMID: 20483874.

22. Piao BK, Wang YX, Xie GR, Mannmann U, Matthes H, Beuth J, Lin HS. Impact of complementary mistletoe extract treatment on quality of life in breast, ovarian and non-small cell lung cancer patients. A prospective randomized controlled clinical trial. Anticancer Research, 2004;24:303–310

23. Tröger W, Zdrale Z, Tisma N, Matijasevic M. Additional therapy with a mistletoe product during adjuvant chemotherapy of breast cancer patients improves quality of life: an open randomized clinical pilot trial. ECAM, 2014;2014:01

24. Thies A, Dautel P, Meyer A, Pfüller U, Schumacher U. Low-dose mistletoe lectin-I reduces melanoma growth and spread in a scid mouse xenograft model. Br J Cancer. 2008 Jan 15;98(1):106–12. doi: 10.1038/sj.bjc.6604106. Epub 2007 Nov 20. PMID: 18026191; PMCID: PMC2359693.

25. Büssing A, Suzart K, Bergmann J, Pfüller U, Schietzel M, Schweizer K. Induction of apoptosis in human lymphocytes treated with Viscum album L. is mediated by the mistletoe lectins. Cancer Lett. 1996 Jan 19;99(1):59–72. doi: 10.1016/0304-3835(95)04038-2. PMID: 8564930.

26. Elluru S, Duong Van Huyen JP, Delignat S, Prost F, Bayry J, Kazatchkine MD, Kaveri SV. Molecular mechanisms underlying the immunomodulatory effects of mistletoe (Viscum album L.) extracts Iscador. Arzneimittelforschung. 2006 Jun;56(6A):461–6. doi: 10.1055/s-0031-1296813. PMID: 16927527.

27. Samtleben R, Hajto T, Hostanska K, et al.: Mistletoe lectins as immunostimulants (chemistry, pharmacology and clinic). In: Wagner H, ed.: Immunomodulatory Agents from Plants. Birkhauser Verlag, 1999, pp 223–41.

28. Hajto T, Hostanska K, Frei K, Rordorf C, Gabius HJ. Increased secretion of tumor necrosis factors alpha, interleukin 1, and interleukin 6 by human mononuclear cells exposed to beta-galactoside-specific lectin from clinically applied mistletoe extract. Cancer Res. 1990 Jun 1;50(11):3322–6. PMID: 2334925.

29. Bussing A. Immune modulation using mistletoe (Viscum album L.) extracts Iscador. Arzneimittelforschung. 2006;56:508–515

30. Tröger W, Jezdić S, Ždrale Z, et al.: Quality of life and neutropenia in patients with early stage breast cancer: a randomized pilot study comparing additional treatment with mistletoe extract to chemotherapy alone. Breast Cancer: Basic and Clinical Research 3: 35–45, 2009.

31. Schad F, Axtner J, Happe A, Breitkreuz T, Paxino C, Gutsch J, Matthes B, Debus M, Kröz M, Spahn G, Riess H, von Laue HB, Matthes H. Network Oncology (NO) – a clinical cancer registry for health services research and the evaluation of integrative therapeutic interventions in anthroposophic medicine. Forsch Komplementmed. 2013;20(5):353–60. Doi: 10.1159/000356204. PMID: 24200825.

32. AbnobaVISCUM®. Summary of product characteristics. Accessed 21.07.2023. https://www.abnoba.de/wp-content/uploads/2022/03/SmPC-all-products_2021_EN-unofficial.pdf

33. Fan Y, Xie W, Huang H, Wang Y, Li G, Geng Y, Hao Y, Zhang Z. Association of Immune Related Adverse Events With Efficacy of Immune Checkpoint Inhibitors and Overall Survival in Cancers: A Systemic Review and Meta-analysis. Front Oncol. 2021 Apr 12;11:633032. doi: 10.3389/fonc.2021.633032. PMID: 33912454; PMCID: PMC8072154.

34. Schoenfeld DA. Sample-size formula for the proportional-hazards regression model. Biometrics. 1983 Jun;39(2):499–503. PMID: 6354290.

35. R Core Team (2021). R: A language and environment for statistical computing. R Foundation for Statistical Computing, Vienna, Austria. URL https://www.R-project.org/.

36. Thomas A. Gerds (2019). prodlim: Product-Limit Estimation for Censored Event History Analysis. R package, version 2019.11.13. https://CRAN.R-project.org/package=prodlim

37. Alboukadel Kassambara, Marcin Kosinski and Przemyslaw Biecek (2021). survminer: Drawing Survival Curves using ‘ggplot2’. R package version 0.4.9. https://CRAN.R-project.org/package=survminer

38. Schad F, Steinmann D, Oei SL, Thronicke A, Grah C. Evaluation of quality of life in lung cancer patients receiving radiation and Viscum album L.: a real-world data study. Radiat Oncol. 2023 Mar 6;18(1):47. doi: 10.1186/s13014-023-02234-3. PMID: 36879290; PMCID: PMC9990362

39. Schad F, Axtner J, Kroz M, Matthes H, Steele ML. Safety of Combined Treatment With Monoclonal Antibodies and Viscum album L Preparations. Integr Cancer Ther. 2018;17:41–51

40. Schad F, Thronicke A. Safety of Combined Targeted and Helixor®Viscum album L. Therapy in Breast and Gynecological Cancer Patients, a Real-World Data Study. Int J Environ Res Public Health. 2023 Jan 31;20(3):2565. doi: 10.3390/ijerph20032565. PMID: 36767928; PMCID: PMC9916034.

41. Thronicke A, Oei SL, Merkle A, Matthes H, Schad F. Clinical Safety of Combined Targeted and Viscum album L. Therapy in Oncological Patients. Medicines (Basel). 2018 Sep 6;5(3):100. doi: 10.3390/medicines5030100. PMID: 30200590; PMCID: PMC6164814.

42. Lin PY, Sun L, Thibodeaux SR, Ludwig SM, Vadlamudi RK, Hurez VJ, et al. B7-H1-dependent sex-related differences in tumor immunity and immunotherapy responses. J Immunol (2010) 185:2747–53. doi: 10.4049/jimmunol.1000496

43. Ye Y, Jing Y, Li L, Mills GB, Diao L, Liu H, Han L. Sex-associated molecular differences for cancer immunotherapy. Nat Commun. 2020 Apr 14;11(1):1779. doi: 10.1038/s41467-020-15679-x. PMID: 32286310; PMCID: PMC7156379.

44. Madala S, Rasul R, Singla K, Sison CP, Seetharamu N, Castellanos MR. Gender Differences and Their Effects on Survival Outcomes in Lung Cancer Patients Treated With PD-1/PD-L1 Checkpoint Inhibitors: A Systematic Review and Meta-Analysis. Clin Oncol (R Coll Radiol). 2022 Dec;34(12):799–809. doi: 10.1016/j.clon.2022.03.010. Epub 2022 Apr 7. PMID: 35400597.

45. Gu Y, Tang YY, Wan JX, Zou JY, Lu CG, Zhu HS, Sheng SY, Wang YF, Liu HC, Yang J, Hong H. Sex difference in the expression of PD-1 of non-small cell lung cancer. Front Immunol. 2022 Oct 20;13:1026214. doi: 10.3389/fimmu.2022.1026214. PMID: 36341395; PMCID: PMC9632486.

46. Leitlinienprogramm Onkologie (Deutsche Krebsgesellschaft, Deutsche Krebshilfe, AWMF): Komplementärmedizin in der Behandlung von onkologischen PatientInnen, Langversion 1.1, 2021, AWMF Registernummer: 032/055OL, https://www.leitlinienprogramm-onkologie.de/leitlinien/komplementaermedizin/

47. Oei SL, Kunc K, Reif M, Weissenstein U, Weiß T, Wüstefeld H, Matthes H, Grah C. Prospective observational stud of advanced or metastatic NSCLC patients treated with Viscum album L. extracts in combination wih PD-1/PD-L1 blockade (PHOENIX-III). Oncol Res Treat, DO 10.1159/000496362, vol. 42, suppl. 2, 10.1159/000496362.

48. Fuller-Shavel, N, Krell, J. Integrative Oncology Approaches to Supporting Immune Checkpoint Inhibitor Treatment of Solid Tumours. Curr Oncol Rep (2024). 10.1007/s11912-023-01492-4

49. Devi S, Grundemann C, Huber R, Kowarschik S. Characterization of Viscum album L. Effect on Immune Escape Proteins PD-L1, PD-L2, and MHC-I in the Prostate, Colon, Lung, and Breast Cancer Cells. Complement Med Res. 2023;30(5):386–392

50. Ma L, Phalke S, Stevigny C, Souard F, Vermijlen D. Mistletoe-Extract Drugs Stimulate Anti-Cancer Vgamma9Vdelta2 T Cells. Cells. 2020;9(6)

51. de Vries NL, van de Haar J, Veninga V, et al. gammadelta T cells are effectors of immunotherapy in cancers with HLA class I defects. Nature. 2023;613(7945):743–750

52. Gemes N, Balog JA, Neuperger P, et al. Single-cell immunophenotyping revealed the association of CD4+ central and CD4+ effector memory T cells linking exacerbating chronic obstructive pulmonary disease and NSCLC. Front Immunol. 2023;14:1297577

53. Nada MH, Wang H, Hussein AJ, Tanaka Y, Morita CT. PD-1 checkpoint blockade enhances adoptive immunotherapy by human Vgamma2Vdelta2 T cells against human prostate cancer. Oncoimmunology. 2021;10(1):1989789

54. Huber R, Classen K, Werner M and Klein R. In vitro immunoreactivity towards lectin-rich or viscotoxin-rich mistletoe (Viscum album L.) extracts Iscador applied to healthy individuals. Arzneimittelforschung, 2006;56:447–456

55. Saha C, Das M, Stephen-Victor E, Friboulet A, Bayry J and Kaveri S V. Differential Effects of Viscum album Preparations on the Maturation and Activation of Human Dendritic Cells and CD4(+) T Cell Responses. Molecules, 2016;21. DOI: 10.3390/molecules21070912

56. Zhou S, Yang H. Immunotherapy resistance in non-small-cell lung cancer: From mechanism to clinical strategies. Front Immunol. 2023 Apr 6;14:1129465. doi: 10.3389/fimmu.2023.1129465. PMID: 37090727; PMCID: PMC10115980.

57. Wang YJ, Fletcher R, Yu J, Zhang L. Immunogenic effects of chemotherapy-induced tumor cell death. Genes Dis. 2018 May 17;5(3):194–203. doi: 10.1016/j.gendis.2018.05.003. PMID: 30320184; PMCID: PMC6176216.

58. Tabiasco J, Pont F, Fournie J J and Vercellone A. Mistletoe viscotoxins increase natural killer cell-mediated cytotoxicity. Eur J Biochem, 2002;269:2591–2600.

59. Fischer A. Charakterisierung der immunmodulatorischen Wirkung von Mistelpräparaten auf Zellen des Immunsystems bei Rindern. Dissertation: Universität Hannover; 2006

60. Büssing A. Viscum album L. – Mechanismen der Zytotoxizität. In: Scheer R, Bauer R, Becker H, Berg AP, Fintelmann V, editors. Die Mistel in der Tumortherapie. Essen: KCV Verlag; 2001. p. 121–134.

61. Kienle G S and Kiene H. Die Mistel in der Onkologie: Fakten und konzeptionelle Grundlagen. Schattauer Verlag; Stuttgart, Germany, 2003

62. Park YR, Jee W, Park SM, Kim SW, Bae H, Jung JH, Kim H, Kim S, Chung JS, Jang HJ. Viscum album Induces Apoptosis by Regulating STAT3 Signaling Pathway in Breast Cancer Cells. Int J Mol Sci. 2023 Jul 26;24(15):11988. doi: 10.3390/ijms241511988. PMID: 37569363; PMCID: PMC10418465.

63. Krysko, D., Garg, A., Kaczmarek, A. et al. Immunogenic cell death and DAMPs in cancer therapy. Nat Rev Cancer 12, 860–875 (2012). 10.1038/nrc3380

64. Schad F, Thronicke A, Hofheinz R-D, Matthes H, Grah C. Patients with Advanced or Metastasised Non-Small-Cell Lung Cancer with Viscum album L. Therapy in Addition to PD-1/PD-L1 Blockade: A Real-World Data Study. Cancers. 2024; 16(8):1609. 10.3390/cancers16081609

65. Hofinger J, Kaesmann L, Buentzel J, Scharpenberg M, Huebner J. Systematic assessment of the influence of quality of studies on mistletoe in cancer care on the results of a meta-analysis on overall survival. J Cancer Res Clin Oncol. 2024 Apr 29;150(4):219. doi: 10.1007/s00432-024-05742-1. PMID: 38679615; PMCID: PMC11056339.

